# Adverse birth outcomes and associated factors among newborns delivered in a western African country: a case‒control study

**DOI:** 10.1101/2022.10.06.22280766

**Authors:** Alexandra Vasconcelos, Swasilanne Sousa, Nelson Bandeira, Marta Alves, Ana Luísa Papoila, Filomena Pereira, Maria Céu Machado

## Abstract

**Background:** Newborns with one or more adverse birth outcomes (ABOs) are at greater risk of mortality or long-term morbidity with health impacts into adulthood. Hence, identifying ABO-associated factors is crucial for devising comprehensive and relevant interventions. The aim of this study was to identify factors that are associated with the occurrence of ABO – prematurity (PTB), low birth weight (LBW), macrosomia, congenital anomalies, asphyxia, and sepsis - among babies delivered at the only hospital of Sao Tome & Principe (STP), a resource-constrained sub-Saharan Western African country.

**Methods:** Hospital-based unmatched case‒control study conducted in STP among newborns from randomly selected mothers from July 2016 to November 2018. Newborns with one or more ABO (gestational age <37 weeks, LBW < 2.5 kg, BW >4 kg, 5-minute Apgar score <7, major congenital anomalies, and probable sepsis based on clinical criteria) were the cases (ABO group), while healthy newborns without ABO were the controls (no-ABO group). Data were collected by a face-to-face interview and abstracted from antenatal pregnancy cards and medical records. Multivariable logistic regression analysis was performed to identify ABO risk factors considering a level of significance α=0.05.

**Results:** A total of 519 newborns (176 with ABO and 343 with no-ABO) were enrolled. The mean gestational age and birth weight of cases and controls were 36 (SD=3.7) weeks with 2659 (SD=881.44) g and 39.6 (SD=1.0) weeks with 3256 (SD=345.83) g, respectively. In a multivariable analysis, twin pregnancy [aOR 4.92, 95% CI 2.25–10.74], prolonged rupture of membranes [aOR 3.43, 95% CI 1.69–6.95], meconium-stained amniotic fluid [aOR 1.59, 95% CI 0.97-2.62], and fewer than eight antenatal care (ANC) visits [aOR 0.33, 95% CI 0.18–0.60] were significantly associated with adverse birth outcomes.

**Conclusion:** Modifiable factors were associated with ABOs in this study and should be considered in cost-effectiveness interventions. The provision of high-quality ANC with eight or more visits should be a priority at ANC service delivery in STP. Twin pregnancies as well as intrapartum factors such as prolonged rupture of membranes and meconium-stained amniotic fluid are red flags for adverse birth outcomes that should receive prompt intervention and follow-up.

## Introduction

Adverse birth outcomes (ABOs) are major global public health problems linked to child mortality and morbidity since they can impact children’s short-and long-term well-being due to neurological and health problems throughout their life course [1,2]. Inevitably, a newborn with an ABO is at a higher risk for mortality than newborns without an ABO [3]. Additionally, ABO may disrupt the family condition, leading to high individual and social costs [4].

The magnitude of ABOs worldwide has dramatically decreased in recent decades, although a large gap still exists between high-income countries and low-and middle-income countries (LMICs), making birth outcomes important measures of health at birth in LMICs [4]. The specific burden of each ABO can vary according to country specificities, although preterm birth is the most well-accepted benchmark for morbidity attributable to early gestation [5]. In recent years, ABOs in LMICs have received attention with a wide range of ABOs being reported across different studies [6-11]. While some studies include indicators for early gestation, such as preterm birth (PTB), fetal growth restriction, low birth weight (LBW) as well as perinatal mortality and fetal loss/miscarriage [4,5], others exclusively analyze the ABO for live newborns at birth [12]. On the other hand, ABO can coexist and share the same underlying risk factors. Etiologies of ABO are complex, multifactorial, physiologically diverse, and not entirely well understood, despite decades of research [5].

Different studies have revealed that diverse risk factors are associated with ABOs [8]. Studies in LMICs have reported numerous sociodemographic factors, maternal factors, previous pregnancy outcomes, neonatal factors, and socioeconomic and health system-related factors [12, 13]. For instance, mothers with previous pregnancy outcomes of PTB or LBW are more likely to have recurrence of these ABO than those without previous PTB or LBW [6,9]. A study on maternal health during pregnancy found that women who had at least one health problem during their pregnancy had a twofold-higher risk of delivering LBW newborns in comparison to women without any health problems (aOR 2.6, CI: 1.4–4.8) [14]. Antepartum problems such as malaria and other infections, anemia, hypertension, hyperglycemia, and obstetric complications are all linked to ABO [9-12]. Lack of adequate ANC, household air pollution from unclean cooking fuels, open defecation, no access to improved water, violence, and other socioeconomic disparities are also considered important risk factors for ABO in sub-Saharan African (SSA) countries [15-17].

Most risk factors contributing to ABO are amenable to modification, although they are not the same across different cultures and socioeconomic statuses within a society [4,18]. Thus, knowing each context-specific reality enables targeting and implementing the most proper evidence-based interventions [3,4].

Sao Tome & Principe (STP) is an LMIC and an SSA country, with limited data on the overall adverse birth outcome at the country level, and in the current era of Sustainable Development Goals (SDG), neonatal mortality remains high, demanding urgent intervention in ABO reduction [19-21]. For this study, ABOs were defined as PTB, LBW, macrosomia, congenital anomaly, birth asphyxia and neonatal sepsis suspicion. This present study is included in a broader project on neonatal health and adverse outcomes [22-24], and the authors studied the determinants for perinatal and neonatal mortality (until the 28^th^ day of life) in STP in another study. This current study aimed to identify the risk factors for adverse birth outcomes among newborns delivered at the only hospital maternity unit in STP.

Knowledge of the burden of adverse birth outcomes and key risk factors will provide policy makers and healthcare practitioners in Sao Tome & Principe with evidence that can be used to inform strategies to achieve reductions in ABO and improve overall perinatal health. Additionally, the findings of this study will help to design targeted interventions and better allocate resources.

## Materials and methods

### Study design

A facility-based unmatched case‒control study was conducted in STP among 519 newborns (176 cases and 343 controls) whose mothers gave birth at Hospital Dr. Ayres de Menezes (HAM) maternity ward.

### Setting

The archipelago of Sao Tome & Principe is a 219 161 inhabitant country, one of the smallest Western SSA countries, with a young population and an annual birth cohort of approximately 6.521 [19]. The rate of deliveries in health units is approximately 98%, with 82.4% occurring at the HAM maternity ward, the only hospital in the country [20,21]. The HAM is a tertiary healthcare facility and receives complicated cases referred from facilities with lower levels of care, as it is the only facility with Comprehensive Emergency Obstetric and Neonatal Care (EmONC) capable of providing blood transfusions and performing cesarean sections.

The maternity unit has a facility-based clinical care unit for ill newborn babies - Newborn Care Unit (NCU) - with six baby cots, which usually receives babies with low Apgar score, prematurity, birth weight <1800 or birth weight >4000 g, congenital malformation, suspected infection, or risk of infection, as well as for observation in case of poor maternal outcome.

It is a resource-constrained NCU, like most units in LMICs, lacking a mechanical ventilator or any continuous positive airway pressure machine, surfactant therapy and enteric feeding for assisting sick babies [25]. The babies receive oxygen through nasal prongs or face masks. In the country, there are only two pediatricians and no neonatologists.

### Participants

The recruitment of newborns’ mothers occurred from July 2016 to November 2018.

The eligibility criteria for participants were as follows: 1) all neonates delivered alive at HAM and 2) newborns who were born outside the hospital but were later admitted at HAM on the day of birth. A total of 535 newborns were initially enrolled.

The exclusion criteria included the following: 1) all neonates delivered at HAM born without any signs of life (stillbirths), 2) newborns whose mothers had cognitive impairment, and 3) adolescent or illiterate mothers who had not obtained permission from their parents or legal guardians to participate in the study. Sixteen met the exclusion criteria (stillbirths), with a total of 519 participants enrolled.

### Selection of cases and controls

Cases (ABO group) were newborns with at least one adverse birth outcome: 1) preterm as defined as a birth that occurred before 37 completed weeks (less than 259 days) of gestation [26]; 2) low birth weight (LBW) as a weight of < 2.5 kg at birth [27,28]; 3) macrosomia as a birth-weight over 4000 g irrespective of gestational age [29]; 4) birth asphyxia as an APGAR score at 5-minutes inferior to seven [30,31].; 5) major congenital anomaly as structural changes in one or more parts of the infant’s body that are present at birth; and/or 6) probable sepsis defined as having an intrapartum infectious risk with neonatal clinical suspicion of early onset sepsis plus admission to NCU for antibiotic treatment.

Controls (no-ABO group) were healthy newborns without adverse birth outcomes (≥ 37 gestational weeks at birth, weight ≥ 2.5 kg at birth and not greater than 4 kg, 5-min APGAR score ≥ 7, no congenital anomaly, and no probable sepsis).

### Sampling method

Sample size followed the WHO-steps approach [32] applying a web-based sample size calculator, Raosoft (http://www.raosoft.com/samplesize.html) which suggested a minimum sample size of S = 355, which placed the right dimension between 355 (95%) and 579 (99%) confidence. A total of 535 participants were enrolled based on the following assumptions: two-sided 95% confidence level, power of 80% to detect an odds ratio of at least 2 for adverse birth outcomes. This sample size was also supported by PASS software (https://www.ncss.com/software/pass/).

Eligible cases were selected through random sampling applied to recruit the newborn’s mother. Each morning, from the pile of mothers’ medical folders, every second interval folder was selected and then carried on asking for consent for enrollment. The study was performed in different months (two weeks every two other months) to avoid seasonal interference (rain season and malaria period), avoiding effects by means of confounding variables by guaranteeing a sample with few biases. Women were invited to participate in the study after admission but were interviewed only after delivery. Invitations were made by the main investigator and occurred during daytime hours, from Monday to Friday. Consenting participants in the sample were followed-up (mother and newborn) throughout their stays until hospital discharge.

### Operational definition

“Adverse birth outcome” in this study implies the presence of at least one or more of the neonatal conditions previously specified in the topic “Selection of cases and controls”. Thus, if the mothers admitted to the labor ward gave birth to a baby with one or more ABOs, then these were considered newborns with an adverse birth outcome (ABO group). Those who gave normal live birth, without the abovementioned adverse birth outcome were labeled “newborns with no-ABO” (no-ABO group) [4]. If newborns first admitted as “newborns with no-ABO” were posteriorly admitted at the Neonatal Unit Care, they were included in the ABO group if one of the abovementioned ABOs were identified.

Gestational age was estimated from the date of onset of the last normal menstrual period or through ultrasound dating of pregnancy.

The definition of birth asphyxia was only determined using the components of the APGAR score table [30,31] since techniques such as umbilical arterial blood gas samples from a clamped section of the umbilical cord are not available in STP. This was similarly applied to both term and preterm infants [23].

Infection as early-onset neonatal sepsis diagnosed in STP is only possible in a suspicion-based algorithm since there are no microbiologic techniques such as, blood culture, available. Therefore, for this study, “probable sepsis” was defined as newborns admitted to NCU due to early suspicious signs and symptoms (hypothermia or fever, lethargy, poor perfusion, prolonged capillary refill time, hypotonia, bulging fontanel, respiratory distress, apnea, and gasping respiration), with a maternal infectious risk and requiring antibiotic intravenous treatment [33,34].

The maternal infectious risk was operationally defined as the sum of all the following risk factors: 1) maternal fever (axillary temperature >37.9 C) at the time of delivery, 2) prolonged rupture of the membrane (≥18 hours), and/or 3) foul-smelling amniotic fluid [35].

### Study variables

The dependent variable was the overall adverse birth outcomes. The independent variables tested in this study included factors grouped into five categories. The first category included the newborns’ maternal sociodemographic factors, such as age, educational status, occupation, marital status, education of the baby’s father, residence, and type of access to water and sanitation. The next category was the preconception main factors, such as women’s pregnancy intention, contraceptive utilization, and current obstetric condition (gravidity, parity, previous abortion, stillbirth, previous cesarean section and preceding birth interval). The third category included ANC service, such as number of visits, gestational age at first ANC visit, obstetric ultrasound, and number of fetuses in the ultrasound (twin pregnancies). This third category also included antepartum factors, such as the presence of medical conditions during the current pregnancy, including maternal anemia (hemoglobin concentration <11 g/dl), bacteriuria, hyperglycemia, malaria, HIV, syphilis, and hepatitis B virus. The fourth category included health facility-related factors (being transferred from another unit and who assisted the delivery) as well as intrapartum factors, such as mode of delivery and complications (fetal malpresentation [36], umbilical cord complication [37], prolonged rupture of membranes [33,38], meconium-stained amniotic fluid [39], postpartum hemorrhage (>500 mL bleeding), preeclampsia (hypertension ≥140/90 mmHg and proteinuria in women who were normotensive at ANC), and obstructed labor [40]). The last or fifth category was the newborn characteristics (gestational age, sex, birth weight) and complications (intrauterine growth restriction, infectious risk, neonatal resuscitation, fetal distress at birth, admission to the NCU and intravenous antibiotic treatment).

Since most pregnant women performed tetanus toxoid vaccination, iron supplementation and blood pressure measurements, these factors were not considered. All mothers stated no consanguinity with the baby’s father and did not have smoking habits; therefore, these risk factors were also not included.

### Data collection

Data on the characteristics of participants were gathered and collected from antenatal pregnancy cards, obstetric maternal and newborn records. For antepartum data, relevant details of the perinatal history and antenatal period were collected systematically from the antenatal pregnancy cards. Intrapartum data were collected from labor follow-up sheets, delivery summaries and maternal medical records. Postpartum data were abstract for newborns from birth charts and/or newborn medical records if admitted to the NCU.

Sociodemographic characteristics were supplemented with a structured administered questionnaire through a face-to-face interview of the mothers 12-24 hours after delivery. This questionnaire was adapted from the STP Demographic and Health Survey (DHS) and other similar studies [18].

### Data quality control

The questionnaires were administered in Portuguese, the national language. The questionnaire was pretested at HAM one month before data collection in 23 mothers, and modifications were made based on the pretest results, mainly adjusting terminology for more culturally-friendly terms. Consent to participate in the study was obtained at the time of admission at HAM, but the interview was held until after the woman was stabilized and ready to be discharged. Continuous follow-up and supervision of data collection were made by the supervisors. The collected data were checked daily for completeness. The principal investigator (a pediatrician) executed and was responsible for the main field activities as follows: 1) obtaining consent and enrollment of the mothers, 2) data collection from ANC cards plus maternal clinical and newborn records, 3) newborns’ clinical exams (for diagnosis confirmation), 4) face-to-face interviews, and 5) to perform all data collection entry into the app survey tool.

### Data management

Data were secured in a confidential and private location, and participants were referred to by identification numbers. The informed consent forms were kept separate from the questionnaires. Both could only be linked by a coding sheet available only to the principal investigator.

### Data analysis

Data were entered into the QuickTapSurvey app (2010-2021 Formstack), an offline survey app tool, and further analyzed using the Statistical Package for the Social Sciences for Windows, version 25.0 (IBM Corp. Released 2017. IBM SPSS Statistics for Windows, Version 25.0. Armonk, NY: IBM Corp.). The captured data were then checked for completeness and accuracy by a qualified biostatistician.

Categorical data are presented as frequencies and percentages or means (standard deviations) and ranges (min-max), as appropriate. In this study, cases were coded as 1, and controls were coded as 0 for analysis. The proportion of missing data ranged from 0.8 to 10% across variables. Missing values higher than 10% were described in the analysis.

The chi-square test was used to compare the proportion of cases and controls between selected categorical variables. To recognize the determinants of ABO, univariable and multivariable logistic regression analyses were performed. In the univariable analysis, explanatory variables with a p value <0.25 were candidates for a multivariable logistic regression model to monitor the influence of confounding variables. With their 95% confidence intervals (95% CI), crude (cOR) and adjusted (aOR) odds ratios were determined to assess the strength and existence of an association. The level of significance α=0.05 was considered.

### Ethics approval and consent to participate

The Ministry of Health of Sao Tome & Principe and the main board of the Hospital Dr. Ayres de Menezes are dedicated ethics oversight bodies, and both approved this study. All methods were performed in accordance with the relevant guidelines and regulations in practice. Written informed consent was obtained from all participants (or their parent or legal guardian in the case of adolescent under 16 or for illiterate pregnant women) after the purpose of the research was explained orally by the principal investigator. Participants’ or their legal representatives’ also consented to have the results of this research work published. Participation in the survey was voluntary, as participants could decline to participate at any time during the study.

## Results

A total of 519 newborns (176 cases and 343 controls) were enrolled. The newborn’s mean gestational age (GA) was 38.73 weeks with a standard deviation (SD) of 2.62 (minimum 25 - maximum 43 weeks). The mean birth weight was 3053.79 ±(SD=649) g (minimum 900 g – maximum 4650 g). Cases had a mean GA and birth weight of 36.02 ±(SD=3.7) and 2659.66 ±(SD=881.44) g, respectively, while their counterparts had 39.61 ±(SD=1.03) weeks and 3256.02 ±(SD=345.83) g, respectively. Mean maternal age was 26.5 years with a standard deviation of 7.03 (minimum 14 - maximum 43 years old). The mean maternal age for cases and controls was 26.99 (SD= ±7.15) and 26.24 (SD= ±6.96), respectively.

### Prevalence of adverse birth outcomes

The current study revealed that 343 (66%) births were healthy live births, while the remaining 176 (34%) were births with child-related adverse birth outcomes. The current study identified 6 types of abnormal birth outcomes (Table 1). Of these, 92 (17.7%) were born preterm, 83 (16%) had low birth weight, and 42 (8.1%) had birth asphyxia. The magnitude of the congenital anomalies was 8 (1.5%). The study also identified a 4% rate of macrosomia and 4% of probable neonatal sepsis.

**Table 1.** Frequency and types of adverse birth outcomes related to the newborns among deliveries attended in HAM, Sao Tome & Principe

| Adverse birth outcomes |  | Frequency | Percent |
| --- | --- | --- | --- |
| Preterm baby or birth | No | 427 | 82.3 |
|  | Yes | 92 | 17.7 |
| Low Birth Weight | No | 436 | 84 |
|  | Yes | 83 | 16 |
| Macrosomia | No | 498 | 96.0 |
|  | Yes | 21 | 4.0 |
| Congenital anomaly | No | 511 | 98.5 |
|  | Yes | 8 | 1.5 |
| Birth asphyxia | No | 477 | 91.9 |
|  | Yes | 42 | 8.1 |
| Sepsis suspicion | No | 498 | 95.9 |
|  | Yes | 21 | 4.0 |
| ABOs observed in a new birth | no ABO | 343 | 66 |
|  | at least 1 ABO | 176 | 34 |
|  | Total | 519 | 100.0 |

Abbreviations: ABOs - adverse birth outcomes.

The maternal characteristics as well as antepartum, intrapartum, and postpartum factors for the total of the participants and for cases versus controls are described in Table 2.

**Table 2.** Maternal characteristics as well as antepartum, intrapartum, and postpartum factors for all participants and for cases (newborns with ABO) versus controls (newborns with no-ABO)

| Variables | TOTAL n= 519 | ABO<br>n= 176<br>n (%) | no-ABO<br>n= 343<br>n (%) | p-value |
| --- | --- | --- | --- | --- |
| **Sociodemographic factors** |  |  |  |  |
| Age |  |  |  | 0.277 |
| 14-16 | 29 (5.6) | 11 (6.3) | 18 (5.2) |  |
| 17-19 | 74 (14.3) | 21 (11.9) | 53 (15.5) |  |
| 20-34 | 330 (63.6) | 108 (61.4) | 222 (64.7) |  |
| ≥35 | 86 (16.6) | 36 (20.5) | 50 (14.6) |  |
| Maternal education |  |  |  | 0.004 |
| none | 19 (3.7) | 7 (4) | 12 (3.5) |  |
| primary | 298 (57.4) | 111 (63.1) | 187 (54.5) |  |
| secondary | 166 (32) | 40 (22.7) | 126 (36.7) |  |
| higher | 36 (6.9) | 18 (10.2) | 18 (5.2) |  |
| Maternal occupation |  |  |  | 0.921 |
| housewife | 326 (62.8) | 111 (63.1) | 215 (62.7) |  |
| student | 36 (6.9) | 11 (6.3) | 25 (7.3) |  |
| employed | 157 (30.3) | 54 (30.7) | 103 (30) |  |
| Marital status |  |  |  | 0.650 |
| union/married | 410 (79) | 137 (77.8) | 273 (79.6) |  |
| single | 109 (21) | 39 (22.2) | 70 (20.4) |  |
| Baby's father education |  |  |  | 0.163 |
| none | 11 (2.1) | 3 (1.7) | 8 (2.3) |  |
| primary | 178 (34.3) | 65 (36.9) | 113 (32.9) |  |
| secondary | 126 (24.3) | 39 (22.2) | 87 (25.4) |  |
| higher | 38 (7.3) | 19 (10.8) | 19 (5.5) |  |
| unknown | 166 (32) | 50 (28.4) | 116 (33.8) |  |
| Residence |  |  |  | 1.000 |
| urban | 231 (45.3) | 77 (45.3) | 154 (45.3) |  |
| rural | 279 (54.7) | 93 (54.7) | 186 (54.7) |  |
| Improved water |  |  |  | 0.917 |
| yes | 377 (72.6) | 127 (72.7) | 250 (72.9) |  |
| no | 142 (27.4) | 49 (27.8) | 93 (27.1) |  |
| Sanitation |  |  |  | 0.516 |
| yes | 272 (52.4) | 96 (54.5) | 176 (51.3) |  |
| open defecation | 247 (47.6) | 80 (45.5) | 167 (48.7) |  |
| **Preconception factors** |  |  |  |  |
| Pregnancy planned |  |  |  | 0.631 |
| yes | 126 (24.3) | 41 (23.3) | 85 (24.8) |  |
| no | 293 (56.5) | 97 (55.1) | 196 (57.1) |  |
| missing | 100 (19.3) | 38 (21.6) | 62 (18.1) |  |
| Ever use of family planning methods |  |  |  | 0.721 |
| yes | 106 (24.6) | 33 (23.2) | 73 (25.3) |  |
| no | 325 (75.4) | 109 (76.8) | 216 (74.7) |  |
| Gravidity |  |  |  | 0.389 |
| 1 | 126 (24.3) | 37 (21) | 89 (25.9) |  |
| 2-5 | 273 (52.6) | 94 (53.4) | 179 (52.2) |  |
| ≥5 | 120 (23.1) | 45 (25.6) | 75 (21.9) |  |
| Parity |  |  |  |  |
| 0 | 153 (29.5) | 50 (28.4) | 103 (30) | 0.622 |
| 1-4 | 316 (60.9) | 106 (60.2) | 210 (61.2) |  |
| ≥5 | 50 (9.6) | 20 (11.4) | 30 (8.7) |  |
| Previous abortion |  |  |  | 0.158 |
| yes | 156 (30.1) | 60 (34.1) | 96 (28.0) |  |
| no | 363 (69.9) | 116 (65.9) | 247 (72.0) |  |
| Previous stillbirth |  |  |  | 0.068 |
| yes | 53 (10.2) | 24 (13.6) | 29 (8.5) |  |
| no | 466 (89.8) | 152 (86.4) | 314 (91.5) |  |
| Poor birth spacing |  |  |  | 0.815 |
| yes | 100 (19.3) | 35 (19.9) | 65 (19.0) |  |
| no | 419 (80.7) | 141 (80.1) | 278 (81) |  |
| Previous caesarean section |  |  |  | 0.592 |
| yes | 15 (2.9) | 6 (3.4) | 9 (2.6) |  |
| no | 504 (97.1) | 170 (96.6) | 334 (97.4) |  |
| ANC |  |  |  |  |
| GA at first ANC visit |  |  |  | 0.835 |
| ≤12 | 272 (62.4) | 91 (61.5) | 181 (62.8) |  |
| >12 | 164 (37.6) | 57 (38.5) | 107 (37.2) |  |
| Number of ANC visits |  |  |  | <0.001 |
| 1-4 | 75 (14.6) | 31 (18.1) | 44 (12.9) |  |
| 5-7 | 237 (46.2) | 101 (59.1) | 136 (39.8) |  |
| ≥ 8 | 201 (39.2) | 39 (22.8) | 162 (47.4) |  |
| Obstetric ultrasound |  |  |  | 0.777 |
| yes | 212 (40.8) | 106 (60.2) | 201 (58.6) |  |
| no | 208 (40.4) | 70 (39.8) | 142 (41.4) |  |
| Twin pregnancy |  |  |  | <0.001 |
| yes | 34 (6.6) | 24 (13.6) | 10 (2.9) |  |
| no | 485 (93.4) | 152 (86.4) | 333 (97.1) |  |
| Maternal anaemia |  |  |  | 0.978 |
| yes | 161 (31) | 54 (30.7) | 107 (31.2) |  |
| no | 240 (46.2) | 81 (46) | 159 (46.4) |  |
| not done | 118 (22.7) | 41 (23.3) | 77 (22.4) |  |
| Bacteriuria |  |  |  | 0.212 |
| yes | 155 (29.9) | 44 (25.0) | 111 (32.4) |  |
| no | 207 (39.9) | 74 (42.0) | 133 (38.8) |  |
| not done | 157 (30.3) | 58 (33.0) | 99 (28.9) |  |
| Hyperglycaemia |  |  |  | 0.538 |
| yes | 16 (3.1) | 7 (4.0) | 9 (2.6) |  |
| no | 346 (66.7) | 113 (64.2) | 233 (67.9) |  |
| not done | 157 (30.3) | 56 (31.8) | 101 (29.4) |  |
| Malaria |  |  |  | 1.000 |
| yes | 3 (0.6) | 1 (0.6) | 2 (0.6) |  |
| no | 385 (74.2) | 131 (74.4) | 254 (74.1) |  |
| not done | 131 (25.2) | 44 (25) | 87 (25.4) |  |
| HIV |  |  |  | 1.000 |
| yes | 3 (0.6) | 1 (0.6) | 2 (0.6) |  |
| no | 486 (99.4) | 164 (99.4) | 322 (99.4) |  |
| Syphilis |  |  |  | 0.340 |
| yes | 5 (1.1) | 3 (1.9) | 2 (0.6) |  |
| no | 463 (98.9) | 154 (98.1) | 309 (99.4) |  |
| HsbAg |  |  |  | 0.810 |
| yes | 16 (3.1) | 4 (2.3) | 12 (3.5) |  |
| no | 300 (57.8) | 103 (58.5) | 197 (57.4) |  |
| not done | 203 (39.1) | 69 (39.2) | 134 (39.1) |  |

| Health facility-related factors |  |  |  |  |
| --- | --- | --- | --- | --- |
| Baby delivered at HAM |  |  |  | 0.592 |
| yes | 504 (97.1) | 170 (96.6) | 334 (97.4) |  |
| no | 15 (2.9) | 6 (3.4) | 9 (2.6) |  |
| Transferred from another unit* |  |  |  | 0.009 |
| yes | 21 (4.0) | 13 (7.4) | 8 (2.3) |  |
| no | 498 (96) | 163 (92.6) | 335 (97.7) |  |
| Delivery assisted by |  |  |  | 0.011 |
| obstetrician | 84 (16.2) | 39 (22.8) | 45 (13.5) |  |
| midwife | 421 (83.4) | 132 (77.2) | 289 (86.5) |  |
| Intrapartum complications and mode of delivery |  |  |  |  |
| Foetal malpresentation |  |  |  | 0.607 |
| yes | 4 (0.8) | 2 (1.1) | 2 (0.6) |  |
| no | 515 (99.2) | 174 (98.9) | 341 (99.4) |  |
| PROM |  |  |  | 0.002 |
| yes | 38 (7.3) | 22 (12.5) | 16 (4.7) |  |
| no | 481 (92.7) | 154 (87.5) | 327 (95.3) |  |
| Pre/Eclampsia |  |  |  | 0.029 |
| yes | 37 (7.1) | 19 (10.8) | 18 (5.2) |  |
| no | 482 (92.9) | 157 (89.2) | 325 (94.8) |  |
| Meconium-stained amniotic fluid |  |  |  | 0.027 |
| yes | 90 (17.3) | 40 (22.7) | 50 (14.6) |  |
| no | 429 (82.7) | 136 (77.3) | 293 (85.4) |  |
| Umbilical cord complication |  |  |  | 0.355 |
| yes | 34 (6.6) | 14 (8.0) | 20 (5.8) |  |
| no | 485 (93.4) | 162 (92) | 323 (94.2) |  |
| Obstructed labour |  |  |  | 0.122 |
| yes | 52 (10) | 23 (13.1) | 29 (8.5) |  |
| no | 467 (90) | 153 (86.9) | 314 (91.5) |  |
| Postpartum haemorrhage |  |  |  | 0.339 |
| yes | 1 (0.2) | 1 (0.6) | 0 |  |
| no | 518 (99.8) | 175 (99.4) | 343 (100) |  |
| Normal Vaginal delivery |  |  |  | 0.034 |
| yes | 433 (83.4) | 138 (78.4) | 295 (86.0) |  |
| no | 86 (16.6) | 38 (21.6) | 48 (14.0) |  |
| Caesarean section |  |  |  | 0.039 |
| yes | 79 (15.2) | 35 (19.9) | 44 (12.8) |  |
| no | 440 (84.8) | 141 (80.1) | 299 (87.2) |  |
| Instrumental vaginal delivery |  |  |  | 0.694 |
| yes | 7 (1.3) | 3 (1.7) | 4 (1.2) |  |
| no | 512 (98.7) | 173 (98.3) | 339 (98.8) |  |

| Newborns characteristics and complications |  |  |  |  |
| --- | --- | --- | --- | --- |
| Gestational Age |  |  |  |  |
| <32 | 16 (3.1) | 16 (9.1) | 0 | <0.001 |
| 32 a 37 | 76 (14.6) | 76 (43.2) | 0 |  |
| 38-41 | 350 (67.4) | 65 (36.9) | 285 (83.1) |  |
| ≥41 | 77 (14.8) | 19 (10.8) | 58 (16.9) |  |
| Sex |  |  |  | 0.308 |
| feminine | 253 (48.7) | 80 (45.5) | 173 (50.4) |  |
| masculine | 266 (51.3) | 96 (54.5) | 170 (49.6) |  |
| Birth weight |  |  |  | <0.001 |
| <1500 g | 17 (3.3) | 17 (9.7) | 0 |  |
| 1500-2499 g | 66 (12.7) | 66 (37.5) | 0 |  |
| 2500 -3999 g | 415 (80) | 72 (40.9) | 343 (100) |  |
| ≥ 4000g | 21 (4.0) | 21 (11.9) | 0 |  |
| IUGR |  |  |  | <0.001 |
| yes | 21 (4.0) | 18 (10.2) | 3 (0.9) |  |
| no | 498 (96) | 158 (89.8) | 340 (99.1) |  |
| Infectious risk |  |  |  | <0.001 |
| yes | 117 (22.5) | 62 (35.2) | 55 (16) |  |
| no | 402 (77.5) | 114 (64.8) | 288 (84) |  |
| Neonatal resuscitation performed |  |  |  | <0.001 |
| yes | 28 (5.4) | 26 (14.8) | 2 (0.6) |  |
| no | 491 (94.6) | 150 (85.2) | 341 (99.4) |  |
| Foetal distress at birth |  |  |  | <0.001 |
| yes | 103 (19.8) | 72 (40.9) | 31 (9.0) |  |
| no | 416 (80.2) | 104 (59.1) | 312 (91.0) |  |
| Admission at NCU |  |  |  | <0.001 |
| yes | 70 (13.5) | 64 (36.4) | 6 (1.7) |  |
| no | 449 (86.5) | 112 (63.6) | 337 (98.3) |  |
| Received antibiotic |  |  |  | <0.001 |
| yes | 112 (21.6) | 72 (40.9) | 40 (11.7) |  |
| no | 407 (78.4) | 104 (59.1) | 303 (88.3) |  |

Bold text for p-value ≤0.05
Abbreviations: GA – gestational age; ANC – antenatal care; PROM – prolonged rupture of membranes; IUGR – intrauterine growth restriction; NCU – neonatal care unit; HAM – Hospital Dr. Ayres de Menezes

### Factors associated with adverse birth outcomes

Univariable binary logistic regression was performed to assess the association of ABOs with different characteristics (Table 3). Crude analysis showed that eight or more ANC visits (cOR 0.34, 95% CI 0.19-0.61) and delivery assisted by a midwife (cOR 0.53, 95% CI 0.33-0.85) were ABO protective factors. Among the intrapartum factors, prolonged rupture of membranes (cOR 2.92, 95% CI 1.49-5.72), preeclampsia (cOR 2.19, 95% CI 1.12-4.28) and meconium-stained amniotic fluid (cOR 1.72, 95% CI 1.09-2.74) were significantly associated with adverse birth outcomes. Regarding the mode of delivery, not having a normal vaginal delivery (cOR 1.69, 95% CI 1.06-2.71) and cesarean section (cOR 1.69, 95% CI 1.04-2.75) were associated with a higher ABO risk. The odds of having ABO were five times higher among twin pregnancies (cOR 5.26, 95% CI 2.45-11.27).

**Table 3.** Factors associated with adverse birth outcomes among newborns delivered at HAM in Sao Tome & Principe

| Variables | ABO<br>n= 176<br>n (%) | no-ABO<br>n= 343<br>n (%) | cOR (95% CI) | p-value | aOR (95% CI) p-value |
| --- | --- | --- | --- | --- | --- |
| **Sociodemographic characteristics** |  |  |  |  |  |
| Maternal education |  |  |  |  |  |
| none | 7 (4) | 12 (3.5) | 1 |  |  |
| primary | 111 (63.1) | 187 (54.5) | 1.018 (0.389-2.661) | 0.972 |  |
| secondary | 40 (22.7) | 126 (36.7) | 0.544 (0.201-1.476) | 0.232 |  |
| higher | 18 (10.2) | 18 (5.2) | 1.71 (0.549-5.351) | 0.353 |  |
| **ANC** |  |  |  |  |  |
| Number of ANC visits |  |  |  |  |  |
| 1-4 | 31 (18.1) | 44 (12.9) | 1 |  |  |
| 5-7 | 101 (59.1) | 136 (39.8) | 1.054 (0.622-1.785) | 0.845 | 1.028 (0.594-1.778), $p=0.922$ |
| $\geq 8$ | 39 (22.8) | 162 (47.4) | 0.342 (0.192-0.609) | $<0.001$ | 0.331 (0.182-0.603), $p<0.001$ |
| Twin pregnancy |  |  |  |  |  |
| yes | 24 (13.6) | 10 (2.9) | 5.258 (2.453-11.268) | $<0.001$ | 4.916 (2.251-10.737) $p<0.001$ |
| no | 152 (86.4) | 333 (97.1) | 1 |  |  |
| **Intrapartum complications** |  |  |  |  |  |
| PROM |  |  |  |  |  |
| yes | 22 (12.5) | 16 (4.7) | 2.92 (1.491-5.716) | 0.002 | 3.429 (1.693-6.946), $p=0.001$ |
| no | 154 (87.5) | 327 (95.3) | 1 |  |  |
| Pre/Eclampsia |  |  |  |  |  |
| yes | 19 (10.8) | 18 (5.2) | 2.185 (1.116-4.280) | 0.023 |  |
| no | 157 (89.2) | 325 (94.8) | 1 |  |  |
| Meconium-stained amniotic fluid |  |  |  |  |  |
| yes | 40 (22.7) | 50 (14.6) | 1.724 (1.085-2.738) | 0.021 | 1.590 (0.966-2.618), $p=0.068$ |
| no | 136 (77.3) | 293 (85.4) | 1 |  |  |
| **Health facility-related factors** |  |  |  |  |  |
| Transferred from another unit* |  |  |  |  |  |
| yes | 13 (7.4) | 8 (2.3) | 3.340 (1.357-8.218) | 0.009 |  |
| no | 163 (92.6) | 335 (97.7) | 1 |  |  |
| Delivery assisted by |  |  |  |  |  |
| obstetrician | 39 (22.8) | 45 (13.5) | 1 |  |  |
| midwife | 132 (77.2) | 289 (86.5) | 0.527 (0.328-0.848) | 0.008 |  |
| Normal Vaginal delivery |  |  |  |  |  |
| yes | 138 (78.4) | 295 (86) | 1 |  |  |
| no | 38 (21.6) | 48 (14.0) | 1.692 (1.056-2.711) | 0.029 |  |
| Cesarean section |  |  |  |  |  |
| yes | 35 (19.9) | 44 (12.8) | 1.687 (1.037-2.745) | 0.035 |  |
| no | 141 (80.1) | 299 (87.2) | 1 |  |  |
| **Newborns complications\*** |  |  |  |  |  |
| IUGR |  |  |  |  |  |
| yes | 18 (10.2) | 3 (0.9) | 12.911 (3.749-44.471) | <0.001 |  |
| no | 158 (89.8) | 340 (99.1) | 1 |  |  |
| Infectious risk |  |  |  |  |  |
| yes | 62 (35.2) | 55 (16.0) | 2.848 (1.866-4.347) | <0.001 |  |
| no | 114 (64.8) | 288 (84.0) | 1 |  |  |
| Neonatal resuscitation |  |  |  |  |  |
| yes | 26 (14.8) | 2 (0.6) | 29.553 (6.926-126.113) | <0.001 |  |
| no | 150 (85.2) | 341 (99.4) | 1 |  |  |
| Fetal distress at birth* |  |  |  |  |  |
| yes | 72 (40.9) | 31 (9.0) | 6.968 (4.329-11.215) | <0.001 |  |
| no | 104 (59.1) | 312 (91.0) | 1 |  |  |
| Admission at NCU |  |  |  |  |  |
| yes | 64 (36.4) | 6 (1.7) | 32.095 (13.531-76.127) | <0.001 |  |
| no | 112 (63.6) | 337 (98.3) | 1 |  |  |
| Received antibiotic |  |  |  |  |  |
| yes | 72 (40.9) | 40 (11.7) | 5.244 (3.357-8.193) | <0.001 |  |
| no | 104 (59.1) | 303 (88.3) | 1 |  |  |

Abbreviations: ANC – antenatal care; PROM – prolonged rupture of membranes; IUGR – intrauterine growth restriction; NCU
– neonatal care unit; cOR: crude odds ratio; aOR – adjusted odds ratio; CI – confidence interval
\* neonatal complications were related as a consequence of the case definition and therefore were not included in the

Newborns with intrauterine growth restriction had thirteen-fold higher odds of ABO (cOR 12.91, 95% CI 3.75-44.47), and newborns with an infectious risk had three-fold higher odds of ABO (cOR 2.85, 95% CI 1.87-4.35).

Performance of neonatal resuscitation (cOR 29.55, 95% CI 6.93-126.11), fetal distress at birth (APGAR score at first-minute inferior to seven) (cOR 6.97, 95% CI 4.33-11.22), admission to the neonatal care unit (cOR 32.10, 95% CI 13.53-76.13) and receiving intravenous antibiotic treatment (cOR 5.24, 95% CI 3.36-8.19) were all related to increased odds of adverse birth outcomes.

Other factors related to the newborn outcomes, such as experiencing fetal distress at birth, needing resuscitation maneuvers, intrauterine growth restriction, being admitted to the neonatal unit care, and receiving antibiotic treatment, were all identified as high-risk for an adverse birth outcome in the univariable analysis. Taking into consideration the definition used to select the cases in this study - PTB, LBW, macrosomia, congenital anomalies, sepsis, and asphyxia – all of the above neonatal factors were related as a consequence of the case definition and therefore were not included in the multivariable model.

### Multivariable model

The variables that were candidates for the multivariable model were number of ANC visits, twin pregnancy, delivery assisted by midwives or obstetricians, preeclampsia, PROM, obstructed labor, cesarean section, meconium-stained amniotic fluid, and infectious risk.

Twin pregnancy (aOR 4.92, 95% CI 2.25-10.74; p<0.001), PROM (aOR 3.43, 95% CI 1.69-6.95, p=0.001), meconium-stained amniotic fluid (aOR 1.59, 95% 0.97-2.62, p=0.068) and fewer than eight ANC visits (aOR 0.33, 95% CI 0.18-0.60, p<0.001) were independently associated with adverse birth outcomes (Table 3).

## Discussion

Assessing neonatal adverse birth outcomes and identifying contributing factors can help avoid neonatal mortality and morbidity thoroughly and thoughtfully. As a result, the goal of this study was to identify the factors related to ABO in neonates admitted at birth to HAM, in the capital city of Sao Tome & Principe. The lack of complete ANC follow-up, multiple births (twins), meconium-stained amniotic fluid and PROM were all identified as significant associated factors for adverse birth outcomes in the current study.

This study showed a rate of 17.7% prematurity and 16% low birth weight, which are higher than the published estimates for Sao Tome & Principe of 12% PTB and 6.6% LBW [2,41,42]. This finding can be related to the fact that gestational age in the country, similar to other studies conducted in LMICs, is mainly estimated based on the last menstrual period [12,42]. Thus, errors in precise date reporting would lead to misclassification of outcomes related to prematurity. Regarding LBW rates in the country, the source used is the UNICEF Multiple Indicator Cluster Surveys (MICS) [20,21]. MICS data rely upon the information provided by the mother about last birth in the previous two years and are associated with a higher risk of recall bias; therefore, these rate discrepancies between this study and official STP data can be linked to the above reasons. The lack of difference between the rate of PTB and LBW in this study is in line with the studies done in Nepal [43], Ethiopia [44] and Kenya [14] as biologically, a preterm baby has a higher risk of having a low birth weight as they are less likely to get sufficient time for maturity, growth, and nutrient intake. Another important consideration is that most PTBs in this study were “late preterm” (76/92), often associated with a normal postnatal clinical course with no relevant complications, when compared to the very preterm babies (16/92) [45].

The 1.5% congenital malformation, 4% macrosomia and 4% neonatal probable sepsis rates found in this study are similar to the rates described by other LMICs [45].

In developing countries, the incidence of neonatal septicemia ranges from 1.6% to 3.8% of all live births, considering that the neonatal sepsis definition varies, and most countries are not able to have a microbiological confirmation, such as STP [46,47]. Moreover, the rates of neonatal sepsis in STP are probably much higher since we only included babies diagnosed while admitted to the maternity unit. Most babies were discharged between the 24^th^ and 36^th^ hours of life; thus, all those with late-onset sepsis were missed from this study.

The 8.1% prevalence of birth asphyxia found in this study is lower when compared to other LMICs: a study in Ethiopia reports a pooled prevalence of 22.52%, 18% in other East African countries and 9.1% in some Central African countries [48]. The lower burden of asphyxia in STP may be due to differences in case definition, as in this study, birth asphyxia was only based on a fifth minute APGAR score less than 7, whereas other studies also use other criteria, such as umbilical cord pH < 7-or 20-min Apgar score less than 7 or multiorgan failure in the first 72 h or convulsion in the first 24 h of life [48]. In turn, APGAR scoring is vulnerable to midwife evaluation and therefore susceptible to higher scoring for better health-related outcomes of the delivery she assisted.

The fivefold higher risk of multiple pregnancies having ABO, can be associated with the fact that monochorionic pregnancies have a vascular anastomosis within the placenta, affecting the perfusion of each twin and promoting adverse outcomes such as preterm labor, premature rupture of membranes, antepartum hemorrhage and fetal death. Adverse outcomes in twin pregnancies were also reported in other studies [49,50]. Some studies describe that twin pregnancies have a thirteen-fold increase in rates of stillbirth in monochorionic pregnancies and a five-fold increase in dichorionic twins compared with singleton pregnancies [51]. Hence, the association between multiple pregnancies and ABO in this study reveals the importance of screening for multiple pregnancies as one key component of ANC to reduce the risk of ABO [49]. Other interventions to reduce ABO related to twin pregnancies that can be recommended in STP are delivery at 37 weeks gestation in uncomplicated dichorionic twin pregnancies and delivery at 36 weeks in monochorionic pregnancies, as proposed by some authors [51].

In this study, complete ANC with eight or more visits was a protective factor, which should be expected, since antenatal care is the most important practice for mothers to obtain more information about nutrition and health, to perform screenings and to learn about danger signs regarding pregnancy and childbirth. If a mother lacks ANC, minor obstetric conditions are not detected and managed early; therefore, serious complications and ABO will likely develop [52]. Our finding is similar to the results from Ethiopia and other LMICs, in which adverse birth outcomes, especially PTB and LBW, were higher among mothers with limited ANC visits [8,18,53]. The recommendation of eight or more ANC contacts was only introduced in 2016 by the World Health Organization (WHO) since studies at that time concluded that eight or more ANC contacts could reduce perinatal deaths by up to 8 per 1000 births when compared to only 4 visits, as previously recommended by the WHO for LMICs [54].

To highlight that this study, in matter of ANC screenings completion, revealed that from all the mothers, a total of 22.7%, 30% and 30% had no screening for anemia, bacteriuria or hyperglycemia done throughout the pregnancy. Therefore, the Ministry of Health of STP should give more attention to expanding antenatal care and create awareness about the importance of a complete ANC follow-up, not only in terms of number of visits but also in terms of completion of all the evidence-based recommended screenings.

The rate of pregnancies with meconium-stained amniotic fluid in this study (17.3%) is similar to the international standard and from other studies in LMICs [10]. The occurrence of meconium-stained amniotic fluid during labor has long been considered a predictor of adverse birth outcomes and an important sign of fetal distress being associated with the rates of neonatal resuscitation, respiratory distress, lower Apgar score, birth asphyxia, neonatal care unit admissions and meconium aspiration syndrome [10,55]. Additionally, approximately 5–10% of neonates with meconium will experience meconium aspiration syndrome, which accounts for approximately 12% of neonatal mortality (as much as a 40% case fatality rate for the neonate and around 2% of perinatal mortality) as well as for neonatal sepsis and pulmonary disease, representing an important risk factor not only for ABO but also for a death outcome [10,55]. Odds of experiencing an ABO, in this study, were identified as being approximately twofold higher among a meconium-stained fluid compared with a clear amniotic fluid at birth. Hence, early detection by using a latent follow-up chart and partograph and timely intervention is recommended to reduce this significant risk factor [10]. Some authors even propose a cesarean section when there is a thick meconium-stained amniotic fluid to ensure a better outcome for the neonate even in the presence of normal fetal heart rate tracings on cardiotocography [55].

PROM was defined as rupture of the membrane lasting more than 18 hours before labor. The overall rate in this study was 7.3%, which is in line with the estimated 5%-10% of all pregnancies in LMICs [56,57]. It was also identified as a significant risk factor for ABO, with a threefold higher risk compared to newborns without PROM. This is a well-known risk factor associated with early onset neonatal sepsis and increased risk for perinatal mortality. Thus, preventive measures should focus on the recognition of these high-risk newborns with early treatment with empirical antibiotics. Such approaches would be a safe and cost-effective strategy, especially in STP where there are no laboratory culture techniques available [56,58].

Studies differ in their observations on risk, and contrary to expectations, no association was found with the sociodemographic maternal characteristics, and there was no relationship between ABO and the maternal sanitation behavior and type of access to water, as reported in other LMICs studies [15,59].

### Strengths and limitations

In this study, the researcher retrieved all clinical pregnancy-related information, namely, ANC screening results, frequency of ANC visits, labor mode of delivery and complications, as well as postpartum care from ANC cards and maternity registers to limit recall bias. The selection of cases and controls was performed by an experienced pediatrician; therefore, it is less likely that this study has misclassification biases both in the exposure and case‒control categories.

Regarding the limitations, maternal factors such as gestational weight gain, BMI, and height were not included in this study, as these measurements are not included in the ANC follow-up in STP, missing key mediators for birth outcomes, as well as strong risk factors for other pregnancy complications [12]. Intrapartum factors, such as antenatal steroids and maternal prophylactic antibiotics, were also not assessed in this study.

The pathogenesis of each of the adverse birth outcomes is multifactorial and we have not identified the specific exposure pathways, such as the incidence of vertical transmission through the birth canal of *group B Streptococcus or Escherichia coli*. Additionally, neonatal sepsis diagnosis was not made by positive culture techniques due to country-resource constraints, which may play a role in our observed outcomes. Previous pregnancy outcomes of PTB or LBW were also not assessed in this study, missing the opportunity to evaluate the recurrence of these ABO compared to those without previous histories of PTB or LBW [6,9].

Notwithstanding these limitations, the study is a first step in the description of adverse birth outcomes among newborns in this resource-constrained setting.

## Conclusions

The high prevalence of adverse birth outcomes in the HAM maternity unit at Sao Tome & Principe could be reduced through the provision of a high-quality ANC and intrapartum care throughout the continuum of care, as fewer than eight ANC visits, twin pregnancy, meconium-stained amniotic fluid and PROM were all identified as significant risk factors for ABOs in this study.

The modifiable risk factors documented in this analysis should be considered in cost-effectiveness interventions that would improve birth outcomes in Sao Tome & Principe. Reducing the ABO burden will not only impact neonatal mortality rates but will also promote child well-being, growth, and favorable health outcomes across their life course and provide substantial population-level human capital returns in Sao Tome & Principe.

## Data Availability

All relevant data are within the manuscript and its Supporting Information files.

## Acknowledgments

A special remark for the late Professor João Luís Baptista PhD MD - AV research cosupervisor - a great man who was a thinker and a fighter for Africa’s improvement of public health. We are indebted to all the women who participated in the study. The authors would like to thank Elizabeth Carvalho and the 1) medical team and nurses of Hospital Dr. Ayres de Menezes Maternity for their support, especially to the chief-nurse Paulina Oliveira, and 2) Ana Sequeira, Rita Coelho, Ana Margalha, Ana Castro, Alexandra Coelho, and Inês Gomes for field support. We would like to acknowledge Instituto Camões, I.P. for the logistic support in Sao Tome & Principe.

